# Homologous and heterologous boosting of the ChAdOx1-S1-S COVID-19 vaccine with the SCB-2019 vaccine candidate: a randomized, observer-blinded, controlled, phase 2 study

**DOI:** 10.1101/2022.05.31.22275010

**Authors:** Sue Ann Costa Clemens, Eveline Pipolo Milan, Eduardo Sprinz, José Cerbino Neto, Filippo Pacciarini, Ping Li, Hui-Ling Chen, Igor Smolenov, Andrew Pollard, Ralf Clemens

**Author notes:** **Corresponding Author** Dr. Ralf Clemens, International Vaccine Institute, SNU Research Park, 1 Gwanak-ro, Gwanak-gu, Seoul, 08826 Korea. **Registered on ClinicalTrials.gov, NCT 05087368**.

## Abstract

**Background:** Ongoing outbreaks of COVID-19 are driven by waning immunity following primary immunizations and emergence of new SARS-CoV-2 variants which escape vaccine-induced neutralizing antibodies. It has been suggested that heterologous boosters could enhance and potentially maintain population immunity.

**Methods:** We assessed immunogenicity and reactogenicity of booster doses of different formulations of alum-adjuvanted SCB-2019 vaccine (9 μg SCB-2019 with or without CpG-1018 adjuvant, or 30 μg SCB-2019 with CpG-1018) in Brazilian adults primed with ChAdOx1-S vector vaccine. S-protein antibodies and ACE2-binding inhibition were measured by ELISA on Days 1, 15 and 29. Participants self-reported solicited adverse events and reactions.

**Results:** All SCB-2019 formulations increased S-protein ELISA antibodies and ACE2 binding inhibition to a greater extent than ChAdOx1-S. After 30 μg SCB-2019+CpG+alum titers against wild-type S-protein were significantly higher than after ChAdOx1-S on Days 15 and 29, as were titers of neutralizing antibodies against wild-type strain and Beta, Gamma, Delta, and Omicron variants. Boosting with SCB-2019 or ChAdOx1-S was well tolerated with no vaccine-related serious or severe adverse events.

**Conclusions:** Boosting ChAdOx1-S-primed adults with SCB-2019 induced higher levels of antibodies against a wild-type strain and SARS-CoV-2 variants than a homologous ChAdOx1-S booster, highest responses being with the 30 μg SCB-2019+CpG+alum formulation.

## INTRODUCTION

The COVID-19 pandemic has been ongoing for two years, during which time large proportions of high-income country populations have achieved vaccine-induced immunity following national immunization campaigns [1]. However, new variants of the wild-type severe acute respiratory syndrome coronavirus 2 (SARS-CoV-2) from Wuhan have continued to emerge each displaying new mutations of the Spike protein (S-protein) [2]. As the S-protein is the main antigenic target of most of the authorized vaccines, the accumulating mutations have resulted in these new variants becoming successively less susceptible to the neutralizing immunity induced by the first immunization campaigns [3–7]. This has resulted in new waves of pandemic COVID-19 outbreaks, most notably associated with the Beta (B.1.351), Gamma (P.1), Delta (B.1.617.2) and Omicron (B.1.1.529) variants [8]. The efficacy of authorized vaccines against infection has been seen to decrease with each new variant, both due to waning immunity following immunization and the changes in the antigenic target, while protection against severe disease is largely preserved.

This has led to the implementation of further immunization campaigns with booster doses of vaccines to broaden the immune response. Early indications are that heterologous boosters are mostly more effective than homologous boosters [9–11]. Most data on such immunity has been obtained with the mRNA vaccines which were the first to be authorized for use in immunization campaigns and have been widely used in high-income countries. However, many low- and middle-income countries are still in the phase of implementing full primary immunization of their populations, with widespread use of viral vector (e.g., ChAdOx1-S1; AstraZeneca, United Kingdom) or inactivated (e.g., CoronaVac; Sinovac Biotech, China) vaccines. Clover Biopharmaceuticals has developed a recombinant SARS-CoV-2 S-protein vaccine (S-Trimer), SCB-2019, that has been stabilized in the native pre-fusion trimeric conformation using the company’s proprietary Trimer-Tag^©^ technology and adjuvanted with the toll-like receptor 9 (TLR 9) agonist CpG-1018 and alum. The SPECTRA phase 2/3 efficacy trial demonstrated that two 30 μg doses of SCB-2019 had 67.2% (95.72% CI: 54.3– 76.8) efficacy against any COVID-19, and specific efficacies (with 95% CI) of 78.7% (57.3– 90.4), 91.8% (44.9–99.8) and 58.6% (13.3–81.5) against Delta, Gamma and Mu variants, respectively [12]. The present study was conducted to investigate use of SCB-2019 in heterologous booster regimens compared with homologous boosters in individuals who have received a two-dose primary vaccination series of the adenovirus-vector vaccine, ChAdOx1-S, which was authorized in Brazil. We assessed safety and immunogenicity of different formulations of SCB-2019; low dose (9 μg) SCB-2019 and alum, with and without the CpG-1018 adjuvant to investigate possible dose-sparing, and the standard 30 μg dose formulated with CpG-1018 and alum used in the SPECTRA efficacy trial. These were given as a heterologous booster in persons primed with two doses of ChAdOx1-S and responses compared with a dose of ChAdOx1-S given as a homologous booster.

## METHODS

This phase 2 randomized, controlled, observer-blinded, multi-center study is ongoing at three sites in Brazil: Hospital de Clínicas de Porto Alegre, Hospital Gloria D’or, Rio de Janeiro and Centro de Estudos e Pesquisa em Moléstias Infecciosas (CEPCLIN), Natal. The study protocol was approved by each hospital’s Ethical Review committee and conducted according to the Declaration of Helsinki and Council for International Organizations of Medical Sciences International ethical guidelines and ICH GCP guidelines. The protocol was registered on ClinicalTrials.gov, registration number NCT 05087368. Participants supplied written informed consent at enrolment. The objective reported here was to select the optimal SCB-2019 formulation to use in the heterologous boosting of individuals primed with two doses of ChAdOx1-S1-S vaccine. Selection was to be based on the safety and immunogenicity, and potential impact on supply of the chosen formulation, i.e. dose-sparing.

### Participants

Eligible participants were male or female adults, ≥ 18 years of age who had previously received two doses of ChAdOx1-S1-S vaccine 6 months (± 4 weeks) before enrolment and were willing and able to comply with study requirements, including all scheduled visits, vaccinations, laboratory tests, and other study procedures. Inclusion criteria included being healthy or having a pre-existing but stable medical condition at the screening examination, the main exclusion criterion being any previous laboratory-confirmed SARS-CoV-2 infection.

### Vaccine

The investigational SCB-2019 vaccine was supplied in a 1.0·mL pre-filled syringe containing 720 µg SCB-2019. Adjuvants were CpG-1018 (Dynavax Technologies) presented in a 2·0 ml vial containing 12 mg/mL of a 22-mer phosphorothioate oligodeoxynucleotide in Tris buffered saline (24 mg per vial), and aluminium hydroxide (Alhydrogel®, Croda Health Care) supplied in vials of 10 mg/mL. The final vaccine formulations per dose contained either 30 μg SCB-2019 with 1·5 mg CpG-1018 and 0.75 mg alum in a 0.5 mL volume, as used in the reported efficacy trial [12], 9 μg SCB-2019 with 0.225 mg alum in a 0.15 mL dose, or 9 μg SCB-2019 with 0.45 mg CpG-1018 and 0.225 mg alum in a 0.15 mL volume.

The comparator vaccine (ChAdOx1-S1-S) was Fiocruz COVID-19 vaccine (Rio de Janeiro, Brazil) containing a Chimpanzee Adenovirus (ChadOx1) encoding the SARS-CoV-2 Spike glycoprotein, with not less than 2.5 × 10^8^ infectious units (Inf.U) in each 0.5 ml dose. These vaccine formulations were prepared on the day of use by trained unblinded vaccine administrators who administered them by intramuscular injection in the upper deltoid of the non-dominant arm. For accurate administration of the 0.15 mL volume 1 mL tuberculin syringes (Precisionglide, Becton Dickenson) were used. These vaccine administrators played no further part in the study and all other study staff and the investigators and participants were masked to which vaccine had been given.

### Procedures

At enrolment participants were randomly allocated 1:1:1:1 using a block size of 8 to four equal groups to receive a third dose of ChAdOx1-S1 or a dose of one of the three formulations of SCB-2019. At their first study visit on Day 1, after providing their baseline blood sample, participants received their assigned vaccination and were monitored for 30 minutes for any immediate reactions. Using diary cards they then recorded for 7 days the occurrence of solicited local reactions (pain, erythema and swelling at the injection site) and systemic adverse events (fatigue, headache, myalgia, arthralgia, loss of appetite, nausea, chills, fever [axillary temperature ≥ 38°C]) daily, together with any unsolicited adverse events, serious adverse events (SAE) or medically attended adverse events (MAAE) occurring up to the third study visit on Day 29. At this study visit the investigator assessed any reported AE as mild (no interference with daily activities), moderate (interferes with daily activities) or severe (prevents daily activity) and the relationship to the study procedures.

### Immunogenicity

Serum samples obtained on Day 1 (before vaccination) and at the second (Day 15) and third (Day 29) visits were used to assess immune responses. The primary endpoint was the response assessed by ELISA as IgG antibodies against SCB-2019 S-protein on Days 15 [12]. Inhibition of binding of S-protein to the human angiotensin converting enzyme 2 (ACE2) was measured using a competitive ELISA with SCB-2019. Additional exploratory analyses included virus neutralizing activity (VNA) titers measured on Days 1, 15 and 29 in a microneutralization assay (MN_50_) against the prototype Wuhan strain and the Beta (B.1.351), Delta (B.1.617.2), Gamma (P.1) and Omicron (B.1.1.529) variants of SARS-CoV-2.

### Statistics

There was no formal hypothesis tested in this first stage of the study, all results being presented and analyzed descriptively. The sample size was considered adequate for the purposes of down-selection of formulation. The primary immunogenicity endpoint was ELISA antibody titers against SCB-2019 S-protein expressed as geometric mean titers (GMT), geometric mean-fold rise in titers over baseline (GMFR) and seroconversion rates (SCR) on Days 15 and 29 in all participants who received the correct vaccination and had no major protocol deviation reported or suffered a COVID-19 infection prior to blood draw. Seroconversion was defined as a ≥ 4-fold increase in post-vaccination titer in those with a baseline titer above the lower limit of quantitation (LLOQ), or a post-vaccination titer ≥ 4-fold the LLOQ in those with no detectable activity at baseline. Comparisons between groups were made by ANCOVA model with vaccine group as a fixed variable, baseline antibody result and site as covariates. Immunogenicity results against the different SARS-CoV-2 variants are also expressed as GMTs, GMFR and SCR for group and variant. GMTs of neutralizing antibodies against prototype strain ELISA GMTs against SCB-2019 S-protein are presented in IU/mL, GMTs of neutralizing antibodies against variants and ACE2 are expressed in reciprocal units. Safety data is presented descriptively as proportions of groups (percentage) reporting any solicited reaction or adverse event or unsolicited adverse events.

## RESULTS

Recruitment began on November 26, 2021 and the last volunteer was enrolled on March 7, 2022. During this period there was a major outbreak of Omicron infections. Of 144 volunteers screened a total of 120 volunteers were enrolled and randomized to the four study groups (**Figure 1**). Only two enrolled participants did not complete through visit 3; one was lost to follow-up from Group 1 after visit 1 and the second was lost to follow-up from Group 2 after visit 2. All of the remaining 118 randomized participants completed through visit 3, except for one from Group 1 who did not provide blood for immunology assessment.

**Figure 1.**
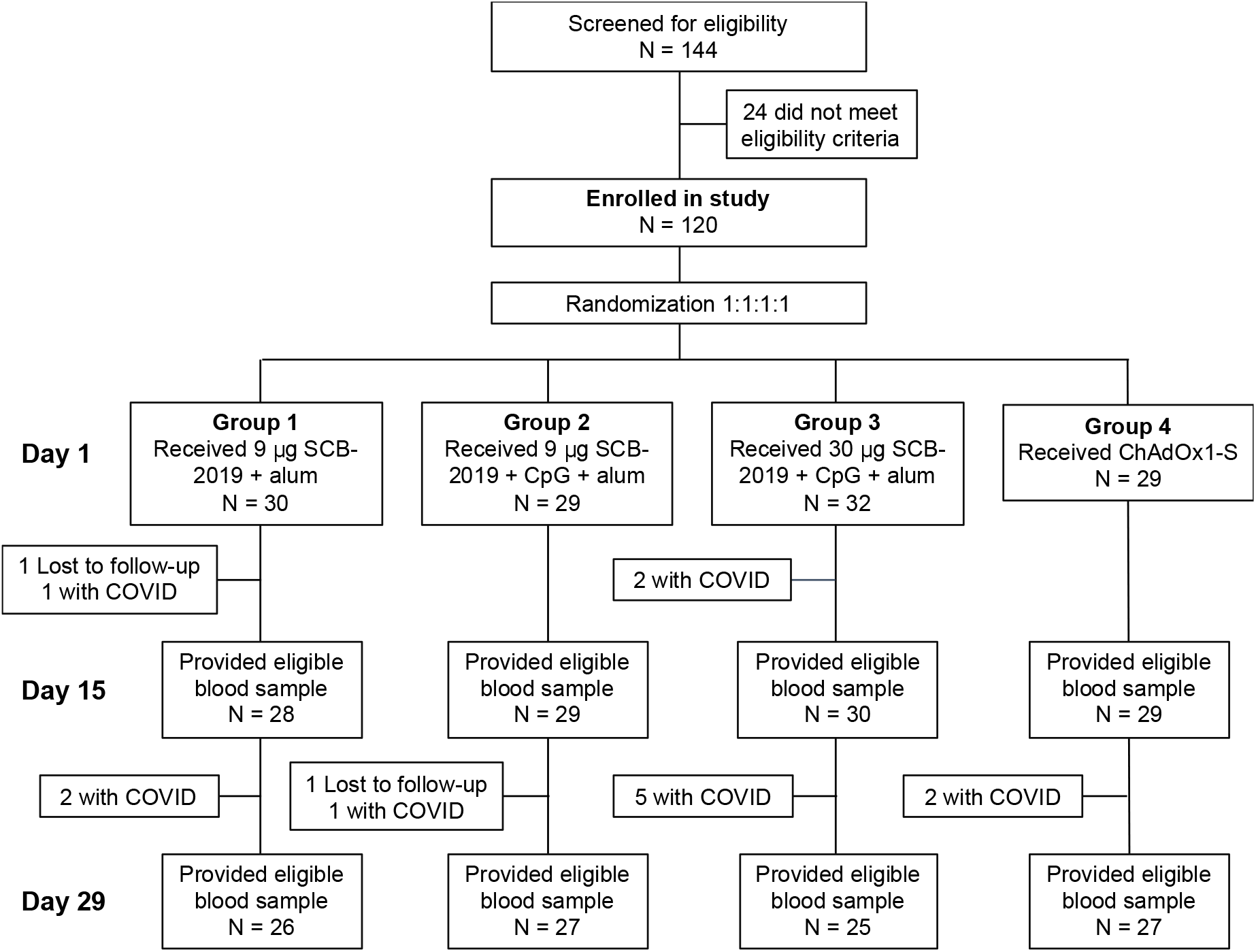
Study flow chart showing the disposition of the participants to each of the four groups.

Demographics in the four study groups were comparable (**Table 1**). Over the course of the study there were 30 suspected cases of COVID-19 in the study population, 26 of which were confirmed by RT-PCR and 2 by rapid antigen test (RAT). These occurred in all four study groups with onset from 8 to 77 days after vaccination (**Table 1)**. Three cases had onset before Day 15, and a further ten before Day 29, the remaining 17 having onset after visit 3. The participants with confirmed COVID-19 infection before the second blood sample were excluded from the relevant immunogenicity analyses.

**Table 1.** Demographics of the participants in the Full Analysis Set (FAS).

| **Characteristics** | **Group 1**<br>9 µg SCB-2019<br>+ alum | **Group 2**<br>9 µg SCB-2019<br>+ CpG + alum | **Group 3**<br>30 µg SCB-2019<br>+ CpG + alum | **Group 4**<br>ChAdOx1-S |
| --- | --- | --- | --- | --- |
|  | N = 30 | N = 29 | N = 32 | N = 29 |
| **Sex, n (%)** |  |  |  |  |
| Male | 15 (50) | 12 (41) | 12 (38) | 13 (45) |
| Female | 15 (50) | 17 (59) | 20 (63) | 16 (55) |
| **Mean age ± SD, yr** | 43.4 ± 14.4 | 40.0 ± 13.6 | 39.8 ± 12.1 | 36.8 ± 12.7 |
| (Range) | (20, 66) | (21, 63) | (22, 63) | (21, 64) |
| **Race, n (%)** |  |  |  |  |
| American Indian/Alaskan native | 0 (0) | 0 (0) | 0 (0) | 1 (3) |
| Black or African American | 1 (3) | 4 (14) | 2 (6) | 4 (14) |
| White | 25 (83) | 22 (76) | 25 (78) | 21 (72) |
| Other | 2 (7) | 3 (10) | 4 (13) | 2 (7) |
| Unknown/not reported | 2 (7) | 0 (0) | 1 (3) | 1 (3) |
| **Ethnic group, n (%)** |  |  |  |  |
| Hispanic or Latino | 21 (70) | 26 (90) | 20 (63) | 19 (66) |
| Not Hispanic or Latino | 2 (7) | 1 (3) | 9 (28) | 4 (14) |
| Unknown/not reported | 7 (23) | 2 (7) | 3 (9) | 6 (21) |
| **Mean body mass index ± SD, kg/m<sup>2</sup>** | 28.5 (4.9) | 28.9 (6.4) | 27.5 (5.6) | 27.8 (6.5) |
| (Range) | (19.8, 37.9) | (17.3, 42.3) | (18.3, 46.3) | (20.0, 44.5) |
| **Risk of severe COVID-19<sup>a</sup>, n (%)** |  |  |  |  |
| Low | 18 (60) | 18 (62) | 24 (75) | 21 (72) |
| High | 12 (40) | 11 (38) | 8 (25) | 8 (28) |
| **COVID-19 infections during the study, n (%)** | 8 (27) | 7 (24) | 9 (28) | 6 (21) |
| Time from vaccination, Mean ± SD, days | 32.4 ± 12.0 | 44.3 ± 14.6 | 24.0 ± 8.2 | 43.2 ± 23.2 |
| (Range) | (8, 47) | (32, 67) | (13, 37) | (8, 47) |

a. Risk due to presence of known co-morbidities

### Immunogenicity

Vaccination in any of the four study groups resulted in increases in titers of binding antibodies against SCB-2019 and inhibition of S-protein binding to ACE2 which were notably higher in Groups 1–3 which received the heterologous SCB-2019 formulation than Group 4 after a homologous ChAdOx1-S booster (**Figure 2**). Following the SCB-2019 boosters these increases were apparently dose dependent; in Groups 1 and 2 which received 9 μg SCB-2019 the respective GMFR from Day 1 were 9 and 11 at Day 15 and 8 and 9 at Day 29, while in Group 3 the GMFR were 18 and 16 at Days 15 and 29 suggesting the 30 μg

**Figure 2.**
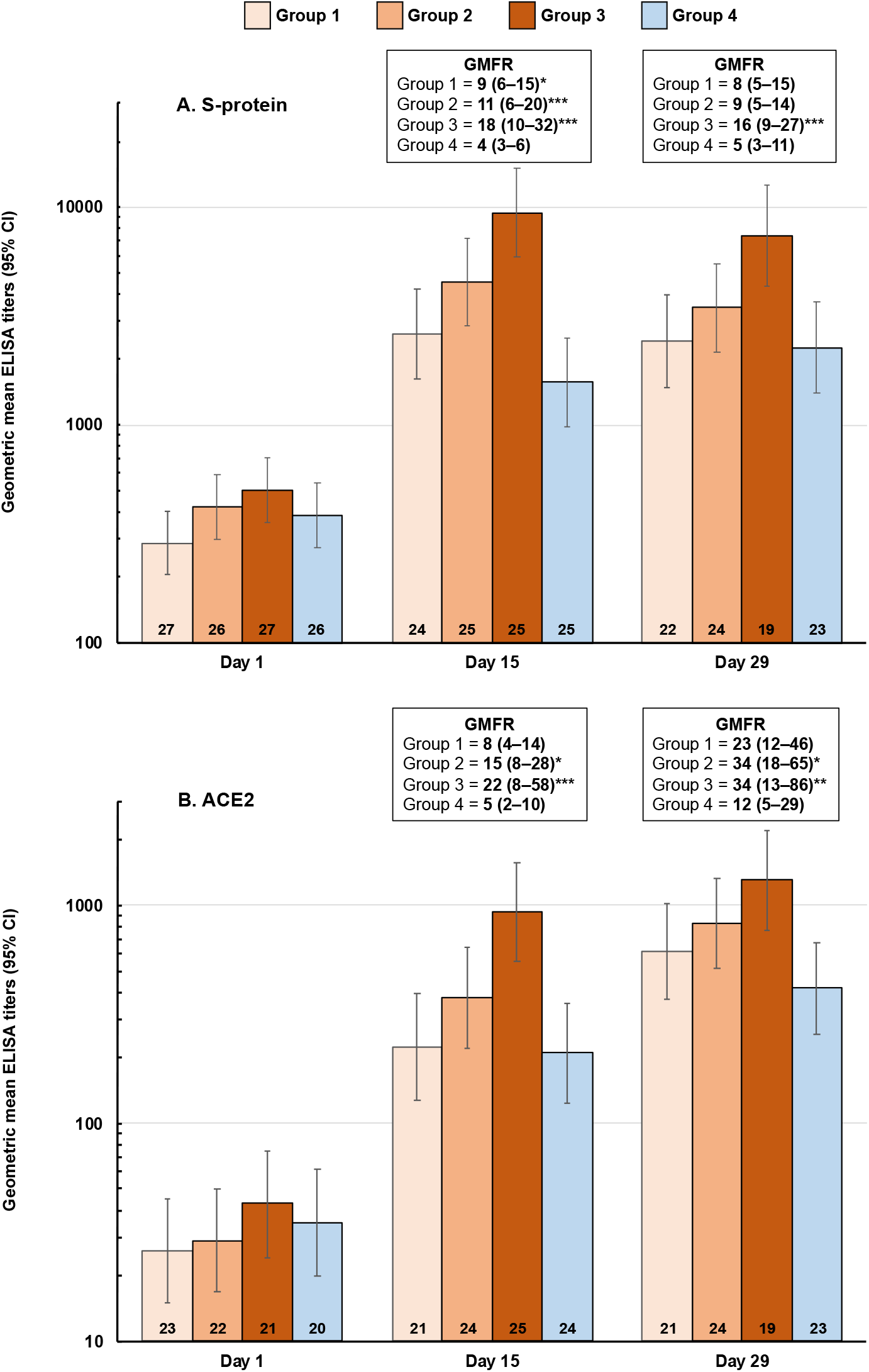
Booster vaccination responses shown as geometric mean titers (95% CI) of ELISA antibodies against SCB-2019 (panel **A**) and ACE2 (panel **B**) at Days 15 and 29 after vaccination. Geometric mean-fold rises (GMFR) from Day 1 (95% CI) are shown with ANCOVA *p* values of differences between Groups 1-3 (SCB-2019) and Group 4 (ChAdOx1-S): * *p* <0.05; ** *p* < 0.01, *** *p* < 0.001). Numbers in columns are n values per group.

SCB-2019 dose was more effective in inducing a booster response (**Figure 2A**). Notably, all three SCB-2019 groups elicited significantly higher GMFR than the four-fold increase elicited by the homologous ChAdOx1-S booster vaccination at Day 15. The response increased further in Group 4 at Day 29, but the significant difference persisted between Groups 3 and 4.

A similar profile was observed when measuring ACE2 binding (**Figure 2B**) with higher fold-increases in all groups at Days 15 and 29 than with the S-protein assay, which were still dose dependent in the SCB-2019 groups. This immune response was significantly higher in Groups 2 and 3 than in Group 4, the ChAdOx1-S-boosted group and this significant difference persisted at Day 29 when al four groups displayed higher titers with GMFR ranging from 23 to 34 in the three SCB-2019 groups compared with a GMFR of 12 in the ChAdOx1-S group.

When assessed for neutralizing activity against the protype strain and the different SARS-CoV-2 variants at Day 15 these differences between different doses of SCB-2019 and between SCB-2019 and ChAdOx1-S were still evident (**Figure 3**). All groups displayed neutralizing activity against all five variants at baseline, the highest responses being against the prototype Wuhan virus and the lowest against the most recent variant, Omicron (B.1.1529). Two weeks after vaccination there were marked increases against all five variants in all four groups, with significantly higher increases after heterologous SCB-2019 doses in Groups 2 and 3 than the homologous ChAdOx1-S dose in Group 4. Overall, the biggest increases in SCB-2019 groups were against prototype virus with GMFR ranging from 10 to 15 (**Table 2**) and Delta variant (GMFR 9 to 17), and lowest against Omicron with GMFR ranging from 4 to 6. The GMFR in Group 4, after a booster ChAdOx1-S vaccination, ranged from 2 to 5 for the different variants. When the GMTs of each group vs Group 4 were compared the responses in Groups 2 and 3 were significantly higher than Group 4 against all variants (**Figure 3**). Similarly, seroconversion rates for each variant were highest with the 30 μg SCB-2019 formulation, with a notably higher response against Omicron than the homologous ChAdOx1-S booster (**Table 2**). Interestingly, the significant differences between Groups 2 and 4, the low-dose fully adjuvanted SCB-2019 and ChAdOx1-S, did into persist to Day 29 except for the Gamma variant. This was due in equal parts to waning titers in Group 2 and increasing titers in Group 4.

**Figure 3.**
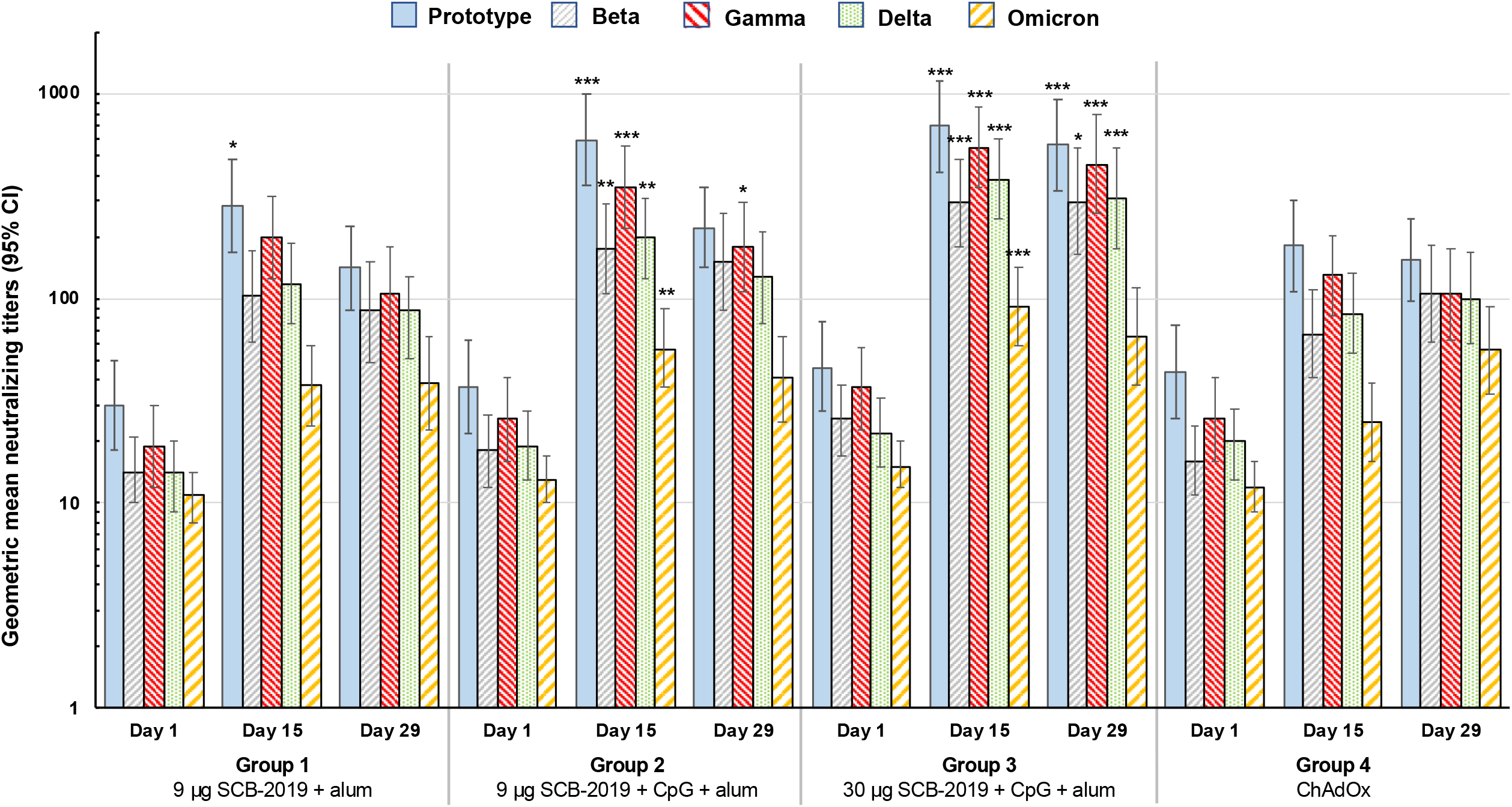
Booster vaccination responses shown as geometric mean neutralizing titers (with 95% CI) against the indicated SARS-CoV-2 variants 15 days after vaccination. Differences in GMTs of Groups 1-3 vs. Group 4 at Day 15 were tested by ANCOVA; * p < 0.05; ** p < 0.01; *** p < 0.001.

**Table 2:** Geometric mean-fold rises (GMFR) at Days 15 and Day 29 from Day 0, and seroconversion rates (SCR) on Days 15 and 29 for antibodies against the prototype SARS-CoV-2 and four variants measured by microneutralization test (MNT_50_).

| Day | Booster vaccine | Group 1<br>9 µg SCIB-2019<br>+ alum | Group 2<br>9 µg SCIB-2019<br>+ CpG + alum | Group 3<br>30 µg SCIB-2019<br>+ CpG + alum | Group 4<br>ChAdOx1-S |
| --- | --- | --- | --- | --- | --- |
| **PROTOTYPE SARS-CoV-2** |  |  |  |  |  |
| **15** | n | 24 | 25 | 25 | 25 |
|  | **GMFR** (95% CI) | **10** (6–17) | **17** (10–30) | **15** (9–27) | **4** (3–7) |
|  | **SCR** (95% CI) | **79%** (58–93) | **84%** (64–96) | **84%** (64–96) | **48%** (28–69) |
| **29** | n | 22 | 24 | 19 | 23 |
|  | **GMFR** (95% CI) | **5** (3–7) | **6** (4–11) | **13** (7–23) | **3** (2–6) |
|  | **SCR** (95% CI) | **64%** (41–83) | **71%** (49–87) | **84%** (60–97) | **39%** (20–62) |
| **BETA variant** |  |  |  |  |  |
| **15** | n | 24 | 25 | 25 | 25 |
|  | **GMFR** (95% CI) | **7** (5–12) | **10** (6–18) | **11** (7–20) | **5** (3–7) |
|  | **SCR** (95% CI) | **75%** (53–90) | **76%** (55–91) | **80%** (59–93) | **52%** (31–72) |
| **29** | n | 22 | 24 | 19 | 23 |
|  | **GMFR** (95% CI) | **6** (4–9) | **10** (6–17) | **12** (7–20) | **6** (3–12) |
|  | **SCR** (95% CI) | **68%** (45–86) | **83%** (63–95) | **90%** (67–99) | **61%** (39–80) |
| **GAMMA variant** |  |  |  |  |  |
| **15** | n | 24 | 25 | 25 | 25 |
|  | **GMFR** (95% CI) | **11** (7–18) | **14** (8–25) | **15** (8–28) | **5** (3–9) |
|  | **SCR** (95% CI) | **79%** (58–93) | **84%** (64–96) | **80%** (59–93) | **56%** (35–76) |
| **29** | n | 22 | 24 | 19 | 23 |
|  | **GMFR** (95% CI) | **6** (4–9) | **8** (5–13) | **13** (7–25) | **4** (2–7) |
|  | **SCR** (95% CI) | **68%** (45–86) | **75%** (53–90) | **79%** (54–94) | **44%** (23–66) |
| **DELTA variant** |  |  |  |  |  |
| **15** | n | 24 | 25 | 25 | 25 |
|  | **GMFR** (95% CI) | **9** (5–14) | **11** (6–19) | **17** (10–27) | **5** (3–8) |
|  | **SCR** (95% CI) | **75%** (53–90) | **76%** (55–91) | **88%** (69–98) | **56%** (35–76) |
| **29** | n | 22 | 24 | 19 | 23 |
|  | **GMFR** (95% CI) | **6** (4–11) | **7** (4–13) | **16** (10–27) | **5** (2–9) |
|  | **SCR** (95% CI) | **68%** (45–86) | **75%** (53–90) | **90%** (67–99) | **48%** (27–69) |
| **OMICRON variant** |  |  |  |  |  |
| **15** | n | 24 | 25 | 25 | 25 |
|  | **GMFR** (95% CI) | **4** (2–6) | **4** (3–7) | **6** (4–10) | **2** (1–4) |
|  | **SCR** (95% CI) | **50%** (29–71) | **64%** (43–82) | **68%** (47–85) | **20%** (6.8–41) |
| **29** | n | 22 | 24 | 19 | 23 |
|  | **GMFR** (95% CI) | **4** (2–6) | **4** (2–6) | **5** (3–8) | **5** (2–9) |
|  | **SCR** (95% CI) | **55%** (32–76) | **58%** (37–78) | **58%** (34–80) | **52%** (31–73) |

Seroconversion defined as a four-fold increase in titer over baseline at Day 1 or from LLoQ if Day 1 titer was < LLoQ.
GMFR = Geometric mean fold rise from Day 1 at Day 15 or Day 29

### Safety

Overall, all four vaccine formulations were well tolerated, with no vaccine-related serious or severe adverse events, no withdrawals due to an AE and no deaths (**Table 3**). The only reported SAE was a leg fracture which was not related to the study procedures. One female participant with a history of hypertension and prior COVID-19 infection reported a mild allergic reaction 13 days after vaccination which resolved but was repeated 5 days later. The investigator considered the first occurrence to be related to the study vaccine, but not the second.

**Table 3.** Reactogenicity in the 29 days after the booster doses of vaccines as indicated in the Safety population.

|  | **Group 1** | **Group 2** | **Group 3** | **Group 4** |
| --- | --- | --- | --- | --- |
| **Vaccine** | 9 µg SCB-2019<br>+ alum | 9 µg SCB-2019<br>+ CpG + alum | 30 µg SCB-2019<br>+ CpG + alum | ChAdOx1-S |
|  | N = 30 | N = 29 | N = 32 | N = 29 |
| **Any solicited local AE** | **8/28 (29%)** | **11/27 (41%)** | **14/30 (47%)** | **9/27 (33%)** |
| Mild | 7/28 (25%) | 11/27 (41%) | 13/30 (43%) | 8/27 (30%) |
| Moderate | 1/28 (4%) | 0/27 (0%) | 1/30 (3%) | 1/27 (4%) |
| **Any solicited systemic AE** | **11/28 (39%)** | **14/27 (52%)** | **13/30 (43%)** | **17/27 (63%)** |
| Mild | 9/28 (32%) | 10/27 (37%) | 7/30 (23%) | 13/27 (48%) |
| Moderate | 2/28 (7%) | 4/27 (15%) | 6/30 (20%) | 4/27 (15%) |
| **Any unsolicited AE** |  |  |  |  |
| Any SOC | 10 (33%) | 19 (66%) | 16 (50%) | 11 (38%) |
| Grade 3 related | 0 | 0 | 0 | 0 |
| Grade 3 not related | 0 | 1 | 0 | 0 |
| **Serious adverse events (SAE)** |  |  |  |  |
| Any | 0 | 1 (3%)* | 0 | 0 |
| Related | 0 | 0 | 0 | 0 |
| **Medically attended AE (MAAE)** | **5 (17%)** | **7 (24%)** | **9 (28%)** | **6 (21%)** |
| **AE of special interest (AESI),**<br>**AE leading to early withdrawal,**<br>**or death** | 0 | 0 | 0 | 0 |

\* One participant suffered a leg fracture which was considered to be unrelated to the study.

There were no clinically meaningful differences in rates of solicited local reactions between the groups. Rates were highest in Group 2 (41%) and Group 3 (47%), after 9 μg and 30 μg SCB-2019 with CpG and alum, respectively. The rate was lower (29%) in Group 1 who received 9 μg SCB-2019 with alum alone. After the homologous ChAdOx1-S vaccination 33% of Group 4 reported a local reaction. Local reactions in all four groups mainly consisted of mild pain at the injection site with a few cases described as moderate but none as severe (**Figure 4**). Solicited systemic AEs were most frequently reported after the ChAdOx1-S in Group 4 (63%), with lower rates (39% to 43%) in the SCB-2019 groups. The most frequent solicited systemic AEs were headache, fatigue and myalgia (**Figure 4**), which were mainly mild and transient, with no severe cases reported. As with local reactions there were no clinically meaningful differences in systemic AEs between groups. There were no AESIs or severe unsolicited AEs reported over the course of the study.

**Figure 4.**
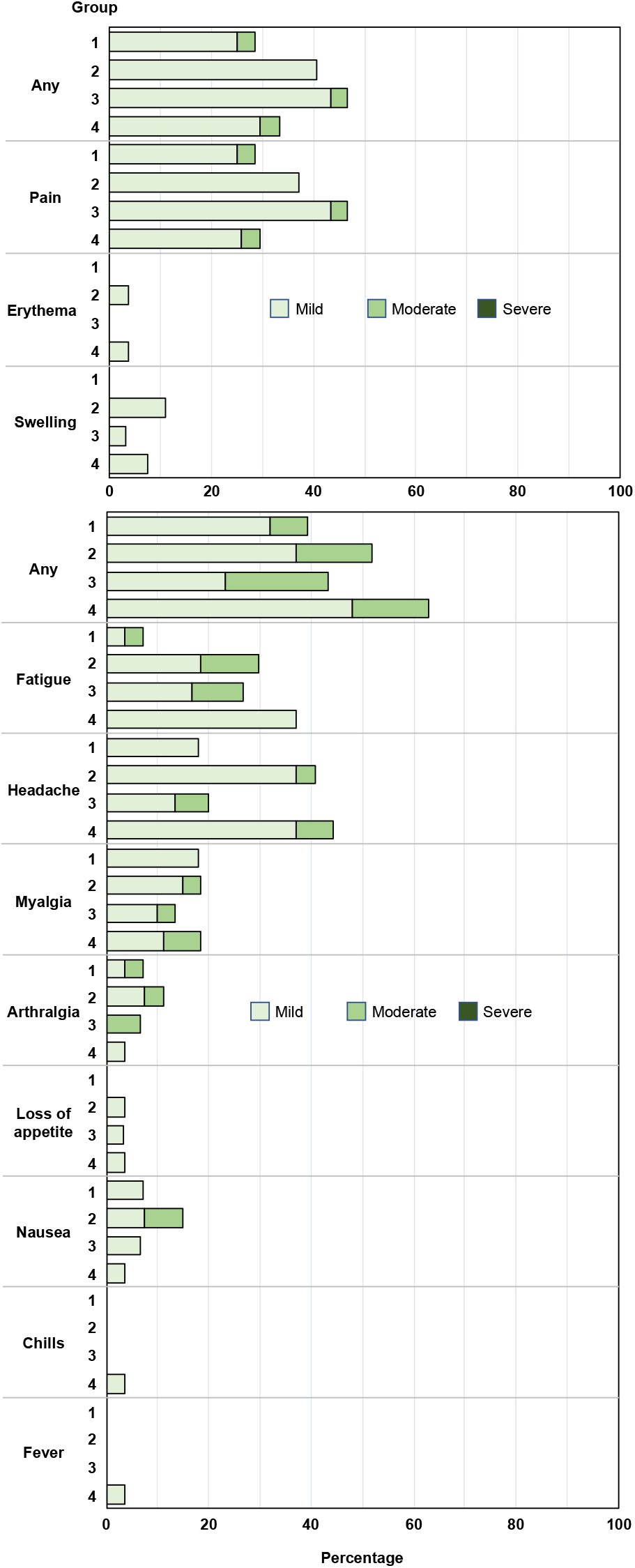
Solicited local reactions and systemic adverse events occurring within 7 days of vaccination by severity reported as percentages of each group.

As noted, there were 30 cases of COVID-19 reported, including 13 from Day 1 to Day 29 when immunogenicity was assessed. Of these 25 were considered to be mild and 5 as moderate in severity; no cases were described as severe or had associated pneumonia or required hospitalization.

## DISCUSSION

This study is the first investigation of boosting immune responses with Clover’s recombinant SARS-CoV-2 S-trimer fusion protein vaccine (SCB-2019) as a heterologous vaccine to the primary vaccine. This study also investigated the use of a lower dose of SCB-2019, with and without the CpG-1018 adjuvant, to allow for dose-sparing. The data show that the best booster response in participants primed with two doses of the adenoviral vector vaccine, ChAdOx1-S, was provided by the standard formulation containing 30 μg SCB-2019 with the toll-like receptor 9 agonist CpG-1018 and alum. This formulation demonstrated 67.2% efficacy against COVID-19 of any severity in the SPECTRA study and 100% efficacy against severe disease [12]. As a heterologous booster it was not associated with any safety concerns and had acceptable reactogenicity, comparable to that observed in the SPECTRA study following primary immunizations [12]. The 30 μg SCB-2019 dose elicited significantly higher immunity against the two key antigenic targets, measured as antibodies against SARS-CoV-2 S-protein and inhibition of the binding of S-protein to the ACE2 receptor, than the homologous ChAdOx1-S vaccine. Further, neutralizing antibody responses against four of the major SARS-CoV-2 variants were significantly higher than with the homologous ChAdOx1-S vaccine. These observations are important as ChAdOx1-S has been shown to be highly protective against severe disease and death since its global roll out.

One exception was the level of neutralizing activity against the Omicron variant; although significantly lower 15 days after homologous boosting than with heterologous with SCB-2019+CpG+alum formulations, titers against Omicron were comparable at Day 29. This was partly due to a continuing increase in these titers in the ChAdOx1-S group while they waned slightly in the SCB-2019 groups. The question of whether this indicates a difference in the kinetics of the response to the homologous booster requires further investigation. It has previously been observed that the immune response to a heterologous second vaccination using mRNA vaccines after a primary dose of ChAdOx1-S is more rapid than the homologous vaccination, but we are unaware of similar observations with a protein or inactivated vaccine [13].

The COVID-19 pandemic has decreased in severity but has endured in numbers of infections with the appearance of new variants which despite appearing to be less sensitive to vaccine-induced immunity, are also leading to less severe forms of disease with fewer hospitalizations and deaths [14]. However, SARS-CoV-2 remains a threat to global health, and the experience of a series of novel variants emerging and rapidly predominating in circulation highlights the potential for future outbreaks. In a situation analogous to influenza, in a population that now has immune experience due to infection or vaccination, future variants may lead to seasonal outbreaks. For that reason, it is essential that high levels of immunity are maintained in global populations to ensure there are no more explosive outbreaks of serious illness such as those the world has recently experienced [15].

Many countries have already achieved high levels of immunity, notably those high-income countries that were able to initiate mass immunization campaigns with the first vaccines to be authorized, mainly the mRNA and vector vaccines targeting the S-protein of the prototype virus [1]. Middle- and low-income countries are now playing catch-up, typically using less expensive and more easily managed inactivated vaccines. However, the steady emergence of a series of novel variants, the majority of which have changes in the main antigenic target, the S-protein, has seen a decline in the extent of protective immunity afforded by the initial vaccines [3–8]. The combination of waning antibodies and lower immunity against the novel variant has caused resurgence of COVID-19 outbreaks around the world, which may be countered by use of booster vaccinations [16]. Unfortunately, boosters only provide a temporary solution as waning immunity and emergence of new escape variants means that the added protection will be short-lived.

Evidence so far suggests that in most cases boosting with a heterologous vaccine is more effective than homologous boosters [17–19] which this report appears to confirm with the heterologous SCB-2019 booster eliciting higher immunity than a dose of the heterologous ChAdOx1-S vaccine. In view of the global need for more COVID-19 vaccines we also assessed the effect of a reduced dose of SCB-2019 to allow dose-sparing as well as omitting the CpG-1018 adjuvant, which may also be dose-limiting. Our results suggest that both formulations of SCB-2019 with CpG and alum containing 9 μg or 30 μg doses, provide an important boost in immunity with no evidence of increased reactogenicity.

This is a small study with several limitations as a consequence, but the trends are confirmation of other observations. Several studies have shown that heterologous booster vaccination can heighten and broaden the immune response compared with homologous booster doses [17–19]. We restricted this study to one priming vaccine, ChAdOx1-S, but results need to be confirmed with other vaccines, particularly mRNA and inactivated vaccines. We only assessed the immune responses out to four weeks after the booster vaccination, and persistence of any improved immune responses following the heterologous and homologous boosters will have to be assessed. Finally, we did not assess the efficacy of the booster immunization; although there were several cases of COVID-19 reported in this small study population it was not designed to include an efficacy assessment which would also require a placebo group. Notably, none of these cases were severe and there were no hospitalizations due to COVID-19.

In conclusion, the formulation of 30 μg SCB-2019 adjuvanted with CpG-1018 and alum is safe and well tolerated and as a heterologous booster vaccine in those previously primed with ChAdOx1-S and is immunologically more effective than that same vaccine given as a homologous booster.

## Data Availability

All data produced in the present study are available upon reasonable request to the authors subject to prior publication of the manuscript in a peer-reviewed journal.

## ACKNOWLEDGEMENTS

The authors wish to thank all participants and our co-investigators Drs. Tannyth Gomes dos Santos, Karla Cristina Marques Afonso Ferreira de Souza, and Deyvison Soares da Costa at CEPCLIN, Natal; Drs. Thaina Garbino, Larisse Longo, and Caroline Portela at Hospital de Clínicas de Porto Alegre, Porto Alegre; and Drs. Bruno Pereira Stuchi, Luciana Gomes Pedro Brandão, and Marcellus Dias da Costa at Hospital Glória D’or, Rio de Janeiro for their invaluable contribution to this study. We are grateful to all the personnel at Vismederi SRL (Siena, Italy) for their expertise in performing the immunogenicity assays, to the staff at Smerud Medical Research (Oslo, Norway) for data management, and to Intrials (São Paulo, Brazil) for their conduct of the study. The ChAdOx1-S vaccine used in the study was generously provided by the Ministry of Health (Brasília, Brazil). The authors are grateful to Keith Veitch (keithveitch communications, Amsterdam, the Netherlands) for assistance with preparation and editorial management of the manuscript.

